# Segmentation and shielding of the most vulnerable members of the population as elements of an exit strategy from COVID-19 lockdown

**DOI:** 10.1101/2020.05.04.20090597

**Authors:** Bram A.D. van Bunnik, Alex L.K. Morgan, Paul R. Bessell, Giles Calder-Gerver, Feifei Zhang, Samuel Haynes, Jordan Ashworth, Shengyuan Zhao, Nicola Rose Cave, Meghan R. Perry, Hannah C. Lepper, Lu Lu, Paul Kellam, Aziz Sheikh, Graham F. Medley, Mark E.J. Woolhouse

## Abstract

In this study we demonstrate that the adoption of a segmenting and shielding (S&S) strategy could increase scope to partially exit COVID-19 lockdown while limiting the risk of an overwhelming second wave of infection.

The S&S strategy has an antecedent in the “cocooning” of infants by immunisation of close family members (Forsyth et al., 2015), and forms a pillar of infection, prevention and control (IPC) strategies (RCN, 2017). We are unaware of it being proposed as a major public health initiative previously.

We illustrate the S&S strategy using a mathematical model that segments the vulnerable population and their closest contacts, the “shielders”. We explore the effects on the epidemic curve of a gradual ramping up of protection for the vulnerable population and a gradual ramping down of restrictions on the non-vulnerable population over a period of 12 weeks after lockdown.

The most important determinants of outcome are: i) post-lockdown transmission rates within the general population segment and between the general and vulnerable segments; ii) the fraction of the population in the vulnerable and shielder segments; iii) adherence with need to be protected; and iv) the extent to which population immunity builds up in all segments.

We explored the effects of extending the duration of lockdown and faster or slower transition to post-lockdown conditions and, most importantly, the trade-off between increased protection of the vulnerable segment and fewer restrictions on the general population.

We illustrate how the potential for the relaxation of restrictions interacts with specific policy objectives. We show that the range of options for relaxation in the general population can be increased by maintaining restrictions on the shielder segment and by intensive routine screening of shielders.

We find that the outcome of any future policy is strongly influenced by the contact matrix between segments and the relationships between physical distancing measures and transmission rates. These relationships are difficult to quantify so close monitoring of the epidemic would be essential during and after the exit from lockdown.

More generally, S&S has potential applications for any infectious disease for which there are defined proportions of the population who cannot be treated or who are at risk of severe outcomes.

## Introduction

As of 3^rd^ May 2020, 3,349,786 confirmed COVID-19 cases and 238,628 COVID-19 related deaths had been reported globally (WHO, 2020). Countries around the world have imposed severe physical distancing measures – ‘lockdown’ – on their entire population to reduce the rate of spread of infection. These measures cause huge (though not fully quantified) societal, psychological and economic harm, and have major indirect impacts on health care provision (OECD, 2020) so there is an urgent need to find ways of exiting lockdown safely.

Here, we consider one option for facilitating exit from lockdown: segmenting and shielding (S&S). Segmenting is dividing the population into groups that are relatively homogeneous in healthcare characteristics or needs (Low et al., 2020). Shielding is a way to protect people who are especially vulnerable to severe COVID-19 outcomes by minimising all interaction between them and other people (British Lung Foundation, 2020).

S&S addresses the concern that while the economic, social and psychological costs of lockdown are distributed across the entire population the public health burden is highly concentrated in identifiable populations of persons “vulnerable” to COVID-19.

Key risk factors for vulnerability to COVID-19 are defined by the World Health Organisation (WHO) as those over 60 years old and those with underlying medical conditions (such as cardiovascular disease, hypertension, diabetes, chronic respiratory disease, and cancer) (WHO, 2020). Although risk factors for severe COVID-19 disease are still incompletely understood, the UK government identified 1.5 million potentially vulnerable individuals who have been advised to shield themselves from infection (Table S1).

There have been numerous mathematical modelling studies of the actual and predicted impact of physical distancing measures on COVID-19 epidemics (e.g. Leung et al., 2020; Bayham et al., 2020; Tuite et al., 2020; Kim et al., 2020; Prem et al., 2020; Block et al., 2020). Very few have explicitly considered shielding (McKeigue and Colhoun, 2020; Neufeld et al., 2020; Weitz et al., 2020) and, despite its inclusion as part of national and international strategy for responding to COVID-19, shielding is not included by any of the mathematical models being used to inform policy in the UK, nor (to the best of our knowledge) any other country. One modelling study in the UK concluded that physical distancing of those over 70 years old (including a 75% reduction of contacts outside home and workplace) would contribute to reducing the burden on the National Health Service (NHS), though lockdown would still be needed to keep burden within NHS capacity (Ferguson et al., 2020).

We therefore constructed a mathematical model designed to explore the complex trade-offs between maintaining or increasing protection for some population segments (shielding) and maintaining or relaxing restrictions on other segments. Key features of our approach include: i) explicit representation of the contact structure between three population segments: vulnerable (v), shielders (s) and the general population (g); and ii) rapidly decaying post-infection immunity.

We use the model to explore the potential of S&S to meet specific policy goals for the UK, namely: i) to save lives; ii) to prevent NHS capacity being overwhelmed; and iii) to protect NHS staff. We consider three, increasingly restrictive, specific objectives that are consistent with these policy goals:

1. future level of infection in the vulnerable population to be kept below the level at the start of lockdown;
2. future levels of infection in the entire population to be kept below levels below levels at the start of lockdown;
3. no increase in numbers of cases or deaths after the start of lockdown.

Objectives (1) and (2) would allow levels of infection to rise in at least some segments at some point in the future. We emphasize that we do not regard any level of infection in any subset of the population as acceptable: COVID-19 can be a serious disease in all age groups and risk groups. However, we suggest that COVID-19 in the non-vulnerable population segments could be managed using a conventional response, centred around good clinical care and proportionate public health measures, without resorting to lockdown of the entire population.

## Methods summary

We developed a susceptible-infectious-resistant-susceptible (SIRS) compartment metapopulation model. Briefly, the population is divided into equal-sized segments with frequency-dependent transmission occurring between segments (see Supplementary Methods for full details). Each segment is comprised of individuals from either the vulnerable, shielder or general population. The contact structure for the baseline realisation of the model is shown in Figure 1.

**Figure 1.**
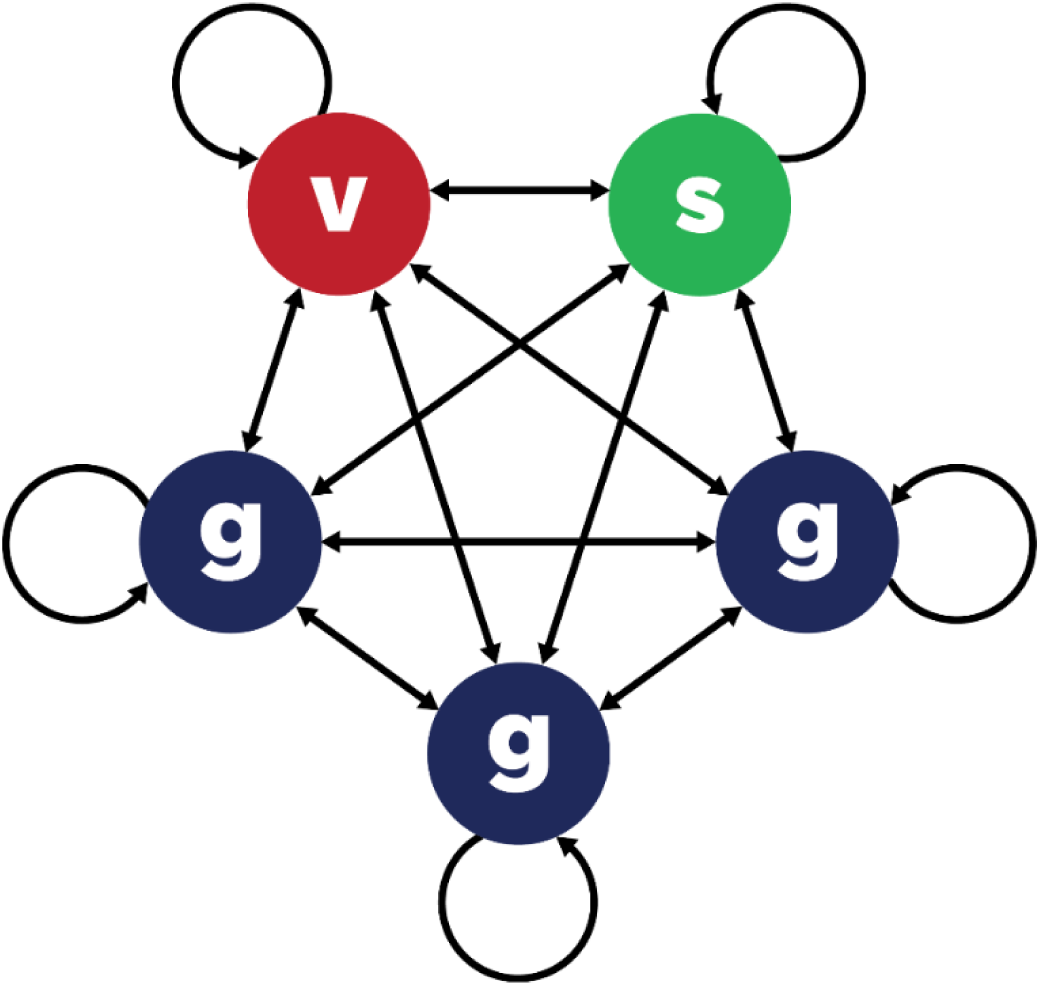
Contact structure for the 20-20-60 model. There are 5 segments, each comprising 20% of the total. v = vulnerable; s = shielders; g = general population. Transmission occurs within and between segments. Transmission rates within and between the three g segments are always homogenous, but may vary within and between segments of different types.

We use the model to explore plausible scenarios for the dynamics of a COVID-19 epidemic during exit from lockdown. We do not make specific predictions; there are too many uncertainties about the epidemiology of COVID-19 for anything other than short-term extrapolations of epidemiological data to be robust. However, we are able to explore the trade-offs that exist between increasing protection for the vulnerable population segments and relaxation of restrictions for non-vulnerable segments. We discuss below how the outputs of the model can be used to inform policy.

Key considerations are the definition of and the size of the vulnerable population. Our approach is informed by public health guidance from the UK government; age and specified underlying health conditions are of primary concern. We therefore consider a set of models including some or all of the following categories:

- individuals >=70 years old (differing from the WHO criterion);
- individuals in receipt of government advice to shield;
- care home residents, those receiving care in the home and hospital patients.

We enumerated these categories using published data (Burton et al., 2019; BLF, 2020; NHS, 2020; ONS, 2019). For our baseline scenario we designated 20% of the total population as vulnerable. We assumed a 1:1 ratio of shielders to vulnerable. The remaining 60% of the population are not in either category and we refer to this as the 20-20-60 model. We estimate that the relative risk of severe disease in the vulnerable 20% is 16:1 (see Supplementary Methods).

We also considered alternative scenarios where the most vulnerable 14%, 8% or 2% are shielded and attributed relative risks of severe disease to these fractions (see Supplementary Methods). We assumed that the smaller the vulnerable population the fewer of their contacts were with the general population: ranging from 3 in 5 for the 20-20-60 model to 1 in 5 for the 2-2-96 model (see Supplementary Methods).

SIRS model parameters were informed by the UK’s Reasonable Worst Case values R_0_=2.8 and doubling time=3.3 days, giving an infectious period of 8.57 days and recovery rate γ=1/8.57 days = 0.117 day^-1^ (National Commissioning Group, 2020).

The contact structures in infectious disease models may be informed by empirical data, e.g. from the POLYMOD study (Mossong et al., 2008). However, such studies cannot inform COVID-19 modelling given the huge impact of physical distancing measures on behaviour. Moreover, the POLYMOD study did not explicitly consider contacts between the vulnerable, shielder and general population segments. We therefore used as simple as possible contact structure that captures the key features of interest here.

Transmission rates, β values, were allowed to vary over four phases (P1-P4). Prior to lockdown (P1) we assumed fully homogenous contact between segments, noting that this implies a force of infection from the general population three times higher than from the vulnerable or shielder populations (Figure 1). We chose β values to give P1 R_e_=1.7 (where R_e_ is the effective reproduction number – see Supplementary Methods for explanation of R_e_), reflecting measures already in place immediately before lockdown, including voluntary self-isolation of cases and quarantining of affected households. During lockdown (P2) we assumed lower values for all β’s including some impact of the shielding advice already in place, giving R_e_=0.8 for the vulnerable population and 0.9 for others. Over a 12-week period after lockdown (P3) we varied β values linearly towards a final value either greater than (relaxation) or less than (protection) P2 values, after which (P4) they remained constant. See Supplementary Methods for full details of β values used.

Initial conditions for the baseline model were chosen to give a cumulative exposure of 6% at t=78 days (one week after start of lockdown), consistent with emerging serological data (PHE, 2020).

We conducted a series of sensitivity analyses on model parameters, including analyses of the impact of different levels of compliance and of active screening of shielders for infection.

## Results

The baseline simulation for the 20-20-60 model generated a scenario in which the combination of increased protection of the vulnerable population and partial relaxation of restrictions for the rest of the population allow a second wave of infection to occur, peaking in the vulnerable population on 141 days after the end of lockdown (Figure 2A). In the vulnerable population the peak was lower than the first peak, but in the other segments it was higher. For this scenario, the percentage of the severe disease burden occurring in the vulnerable population is reduced from 80% to 55% (Table 1).

**Figure 2.**
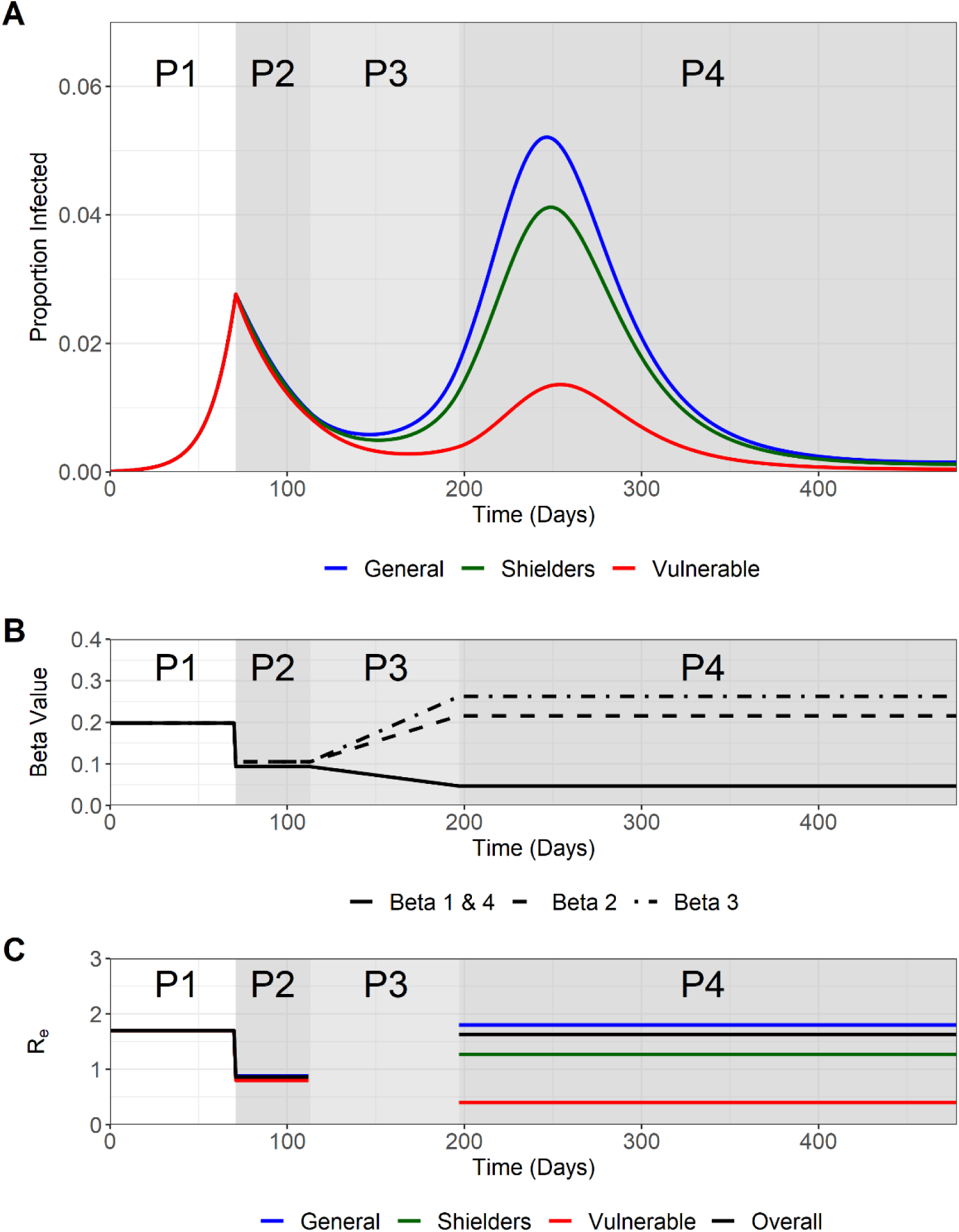
Trajectory plots for the proportion infected in the vulnerable, shielders and general populations, with accompanying β and R_e_ plots. Phases 1-4 are indicated. A) Trajectory plots of the proportion of those infected in the vulnerable (green), shielders (red) and general (blue) populations, shading depicts the different phases of enhanced shielding intervention. B) Values for the different β over the course of the simulation as they are implemented for the different intervention phases. C) Values of the corresponding R_e_ values (colours) for the different subpopulations and the overall R_e_ (black) during the different intervention phases.

**Table 1.** Comparison of the estimated distribution of COVID-19 burden for the 20-20-60, 14-14-72, 8-8-84 and the 2-2-96 scenarios.

| Model | Segment | Proportion of population | Fraction of severe disease burden | Relative risk of severe disease | Cumulative incidence* | Proportion of severe disease burden* |
| --- | --- | --- | --- | --- | --- | --- |
| **20-20-60** | v | 0.20 | 0.80 | 16 | 0.19 | 0.55 |
|  | s + g | 0.80 | 0.20 | 1 | 0.60 | 0.45 |
| **14-14-72** | v | 0.14 | 0.68 | 13.1 | 0.22 | 0.40 |
|  | s + g | 0.86 | 0.32 | 1 | 0.68 | 0.60 |
| **8-8-84** | v | 0.08 | 0.50 | 11.7 | 0.24 | 0.25 |
|  | s + g | 0.92 | 0.50 | 1 | 0.74 | 0.75 |
| **2-2-96** | v | 0.02 | 0.20 | 12.3 | 0.27 | 0.08 |
|  | s + g | 0.98 | 0.80 | 1 | 0.79 | 0.92 |

\*Over one year period from the end of P2 (days 113 to 478).

The modelled changes in β values (Figure 2B) translated into changes in the underlying effective reproduction number, R_e_. For our baseline simulation during Phase 4 although R_e_ was <1 for the vulnerable population it was >1 in both non-vulnerable segments (highest in the general population) and overall (Figure 2C). This has two implications. Firstly, that outbreaks in the vulnerable population are self-limiting and, secondly, that the eventual decline in the epidemic is due to the build-up of population immunity (Figure S1). We note that P2 R_e_<1 implies that if lockdown were continued then levels of infection in all segments would eventually fall to very low levels.

Extending P2 beyond 6 weeks resulted in peaks that were delayed (by more than the extension to the lockdown) but were slightly higher (Figure S2). Extending or shortening Phase 3 by ±6 weeks resulted in peaks that were 37 days later or 37 days earlier respectively but were of similar magnitude (Figure S3).

Varying the start of P2 relative to the epidemic curve had a major impact on subsequent dynamics (Figure S4). This reflects substantial differences in the fractions exposed to infection and therefore the build-up of population immunity. Notably, if the lockdown started earlier in the epidemic curve than estimated (lower I(t)) then the risk of an overwhelming second wave is substantially greater (Figure S4A).

Varying P2 β values (and so R_e_) had an effect on epidemic dynamics, not altering the qualitative outcome but substantially affecting numbers of cases in all three subpopulations (Figure S5).

Varying P3/4 β values had a substantial effect on epidemic dynamics, and could alter the outcome. If P4 R_e_ is greater than 1.99 then the second I_v_ peak exceeds the height of the first (Figure 3A).

**Figure 3.**
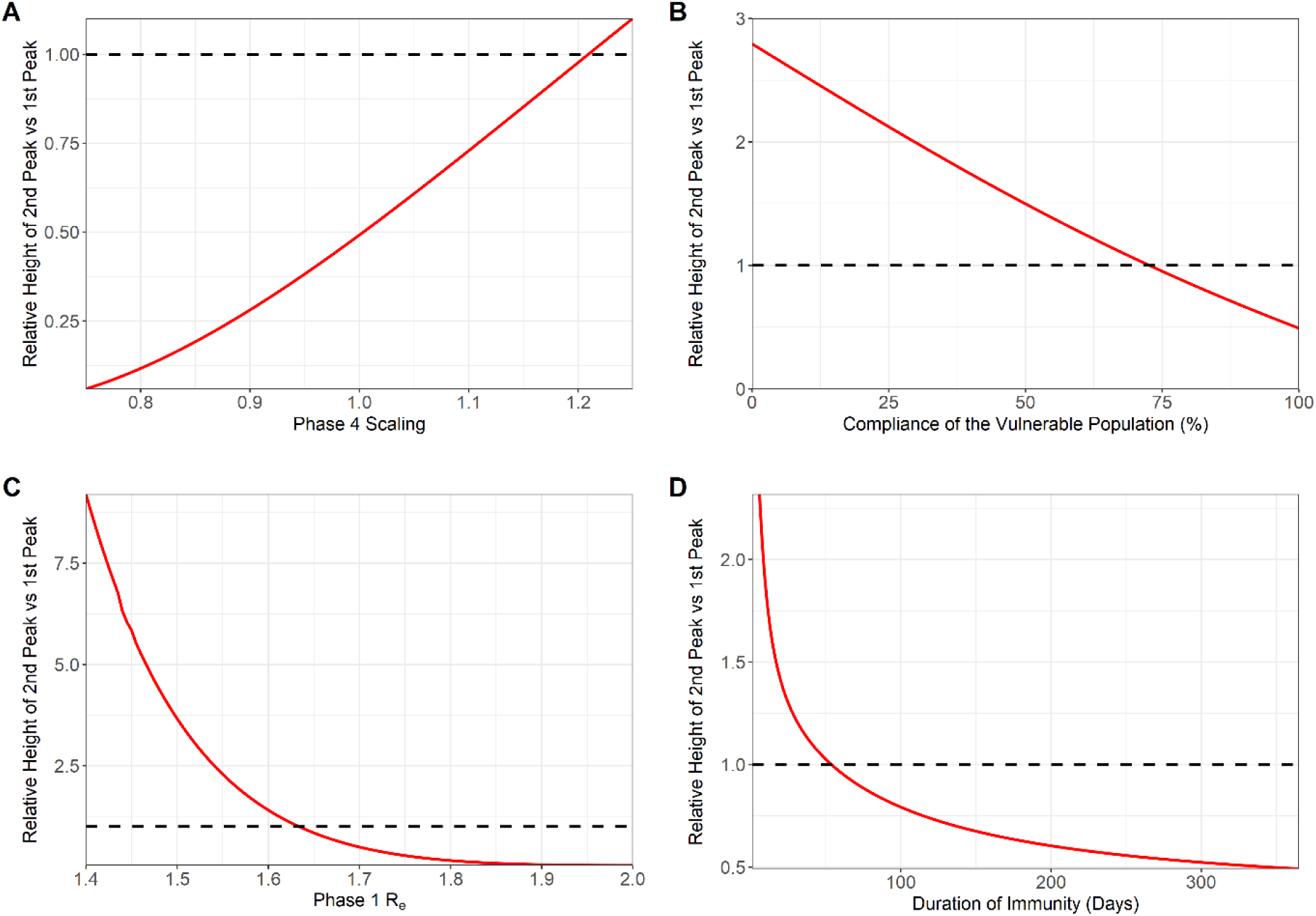
Sensitivity analyses. Plots show the relative height of 2^nd^ peak versus 1^st^ peak I_v_ as a function of relevant parameter value. Dotted lines represent peaks of equal height. A) Relative values of R_e_ in P3/P4. Second peak is higher for relative value >1.22, corresponding to R_e_>1.99. B) Adherence in P3/P4. 100% adherence equates to P4 R_e_=0.4 (baseline value); 0% adherence equates to pre-lockdown value of R_e_=1.7. Second peak is higher for adherence <74%. C) R_e_ in all phases. P1 R_e_ values are shown; R_e_ values in other phases are scaled accordingly. Second peak is higher for P1 R_e_ <1.63. D) Duration of immunity (expressed as 1/ζ). Second peak is higher for 1/ζ <54 days.

Variation in adherence by the vulnerable population during P3/4 was modelled as an impact on β1 and β4 values (Table M3), 100% adherence corresponding to the baseline scenario target values and 0% to a return to Phase 1 values. Assuming that adherence has a linear effect on β1 and β4 values, if adherence is less than 74% then the second I_v_ peak can exceed the height of the first (Figure 3B).

Varying R_e_ throughout also had a significant impact on the outcome. At higher R_e_ values the second peak remained low, but at slightly lower values than our baseline scenario (<1.63 in P1) the second I_v_ peak exceeds the height of the first peak (Figure 3C). This is because a smaller fraction was exposed in the first wave of the epidemic, so there was less population immunity.

Varying the rate of loss of immunity, ζ, also had a significant impact on whether the second peak in the vulnerable population exceeded the first (Figure 3D). At longer average duration of immunity (1/ζ) the second peak remained low, but for shorter durations (<54 days) it exceeds the height of the first peak. This illustrates that epidemic dynamics are highly sensitive to the duration of immunity and its impact on the development of population immunity.

Fourier Amplitude Sensitivity Test (FAST) analysis indicated that key outcomes are differentially sensitive to variation in individual or sets of β values (Figure 4). Three outcome measures were assessed: height of the second peak; whether the second peak is higher than the first; and cumulative incidence over one year. The value of transmission parameters within the general population and between the general and vulnerable populations have the greatest impact on outcomes.

**Figure 4.**
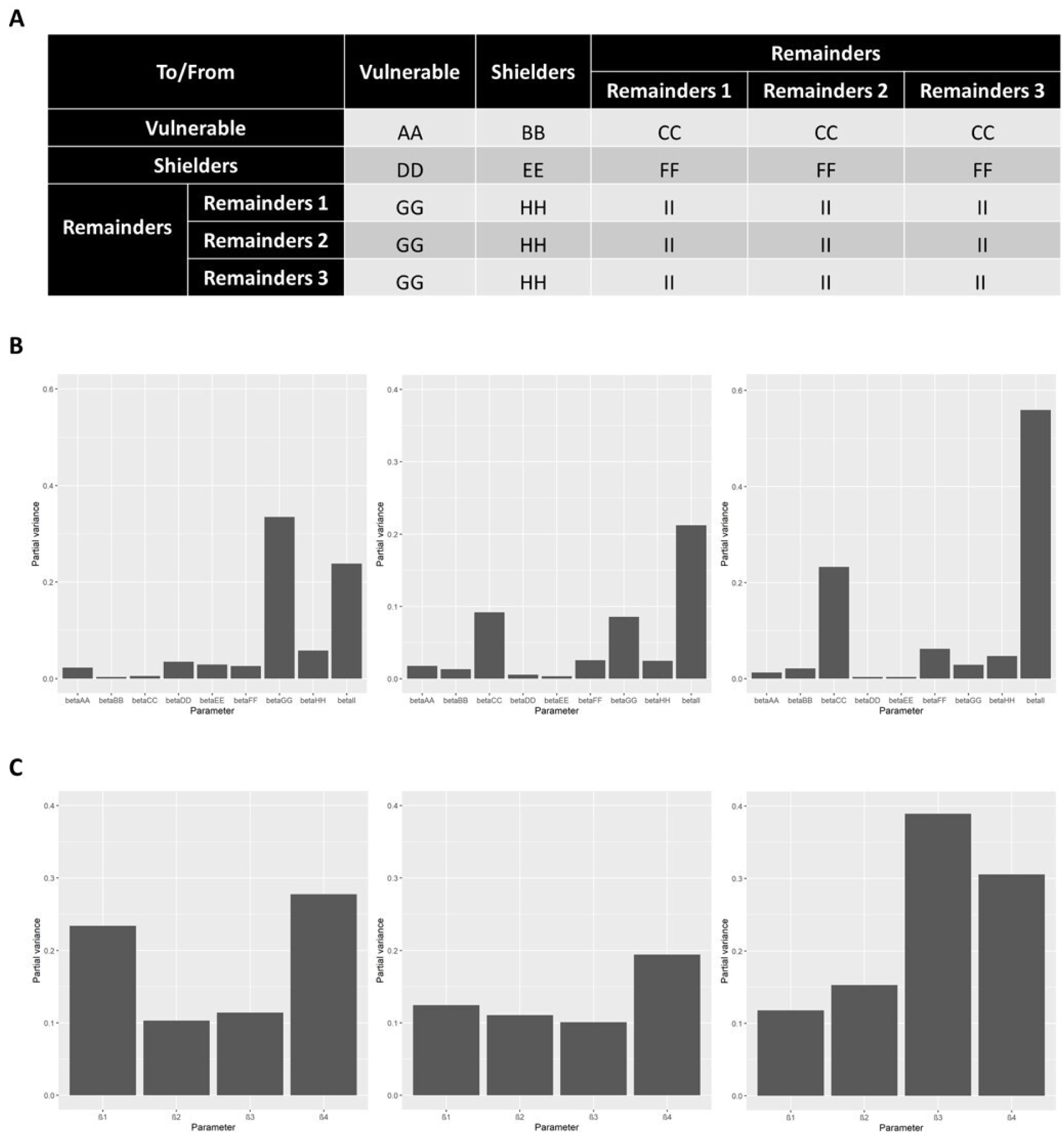
Results of a global sensitivity (FAST) analysis on three key outcome measures with regards the proportion of the vulnerable population that become infected (I_v_) 1) the height of the second peak of I_v_; 2) whether the second peak of I_v_ is higher than the first peak and 3) cumulative I_v_ one year after the start of the lockdown. The bars show the partial variance of the individual model parameters. Higher bars indicate greater sensitivity of the model to that parameter. See Supplementary Methods for details of the sensitivity analysis and parameter ranges used. A) Description of explored β value “blocks” for the sensitivity analysis. β_1_, β_2_, β_3_ and β_4_ were broken down further to assess the sensitivity of the system to these values in greater detail. Lettering denotes the explored β in the FAST analysis. B) Sensitivity of the model outcome measures to the β values specified in A). C) Sensitivity of the model outcome measures to β_1_, β_2_, β_3_ and β_4_.

There is a clear, though asymmetric, trade-off between increasing protection of the vulnerable population and relaxing restrictions on the non-vulnerable population (Figure 5A). This trade-off can be expressed in terms of combinations of protection and relaxation that meet specific policy objectives (Figures 5B-D). The more restrictive the policy objectives (increasing from 5B to 5D) the smaller the parameter space that satisfies those objectives.

**Figure 5.**
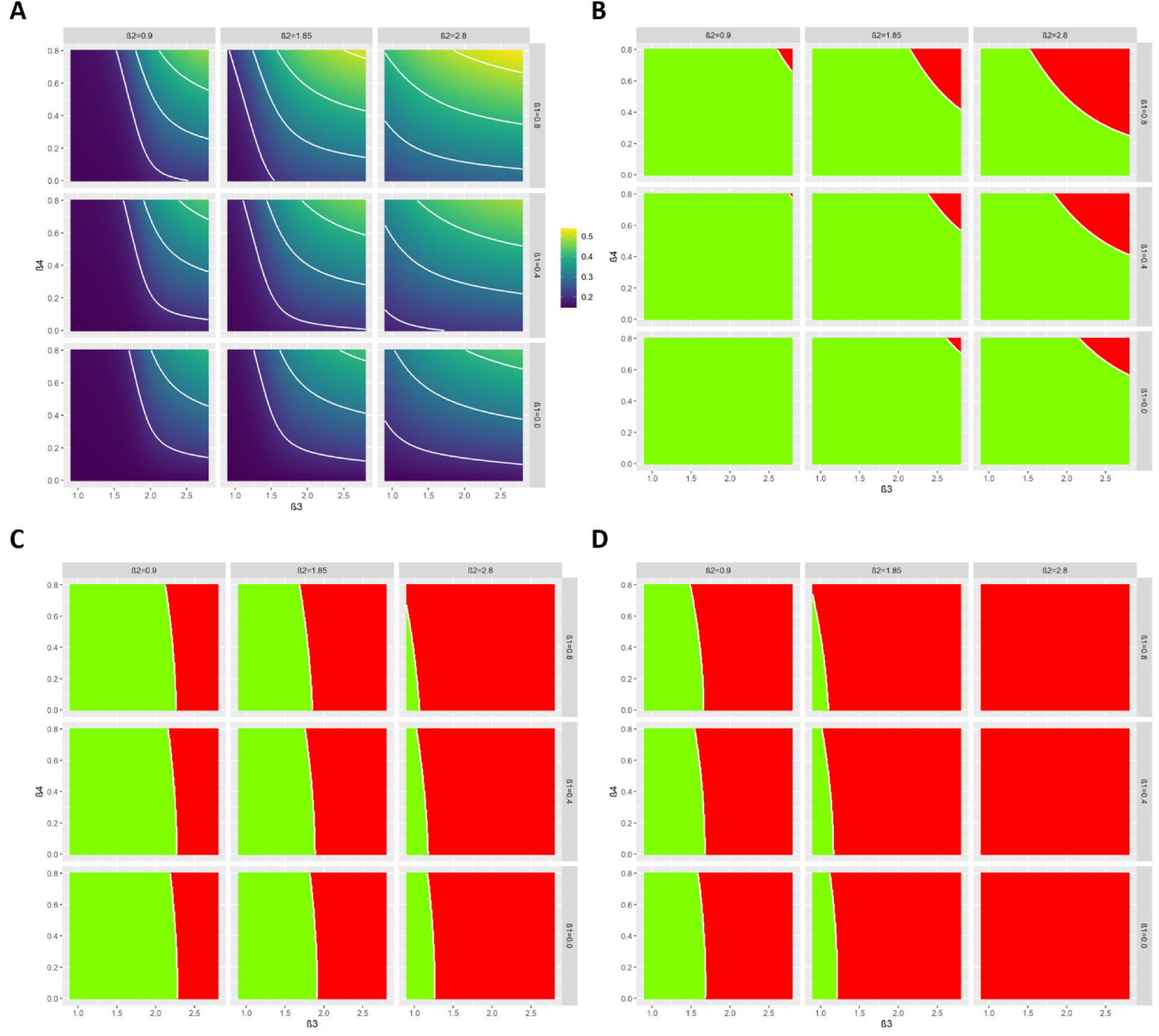
Heat maps showing the trade-off between relaxation (left to right on horizontal axis) and increasing protection (top to bottom on vertical axis). A) Heat maps describing the cumulative infected vulnerable fraction (I_v_) one year after the start of lockdown for different combinations of β_3_ and β_4_ for different values of β_1_ (rows) and β_2_ (columns). B) As A) but for whether the second peak of I_v_ is lower (green) or higher (red) than the first peak. C) As (B) but all 2^nd^ peaks (I_v_, I_s_, I_g_) smaller than 1^st^ peaks (green). D) As (B) but dI/dt is negative or zero for at least 1 year after the start of lockdown for all I-compartments.

The higher the ratio of shielders to vulnerable (taken to be 2:1; 1:1 or 0.5:1) the more the second peaks were delayed and suppressed (Figure S6). This reflects that different fractions of the total population (more or fewer shielders) are subject to greater restrictions.

Moving from the 20-20-60 model to the 14-14-72, 8-8-84 and 2-2-96 models, i.e. decreasing the vulnerable fraction and increasing the proportion of their contacts with shielders, allowed higher and earlier second peaks (Figure S7). This resulted in increased cumulative incidence in both the vulnerable and the shielder plus general population segments (Table 1). At the same time the fraction of the severe disease burden in the vulnerable segment decreased. Together, this makes S&S less effective for narrower definitions of the vulnerable segment.

The 20-20-60, 14-14-72, 8-8-84 and 2-2-96 models generate different trade-offs in terms of combinations of protection and relaxation that meet specified policy objectives (Figure 6). The trade-offs are complex but two key patterns are apparent: as the size of the vulnerable fraction is decreased there is: i) a larger parameter space where no policy objective is satisfied; and ii) much less scope for increasing β3, i.e. the rate of contact within the general population. These constraints can be partially eased by keeping β2 as low as possible, i.e. minimizing contacts between shielders and the general population.

**Figure 6.**
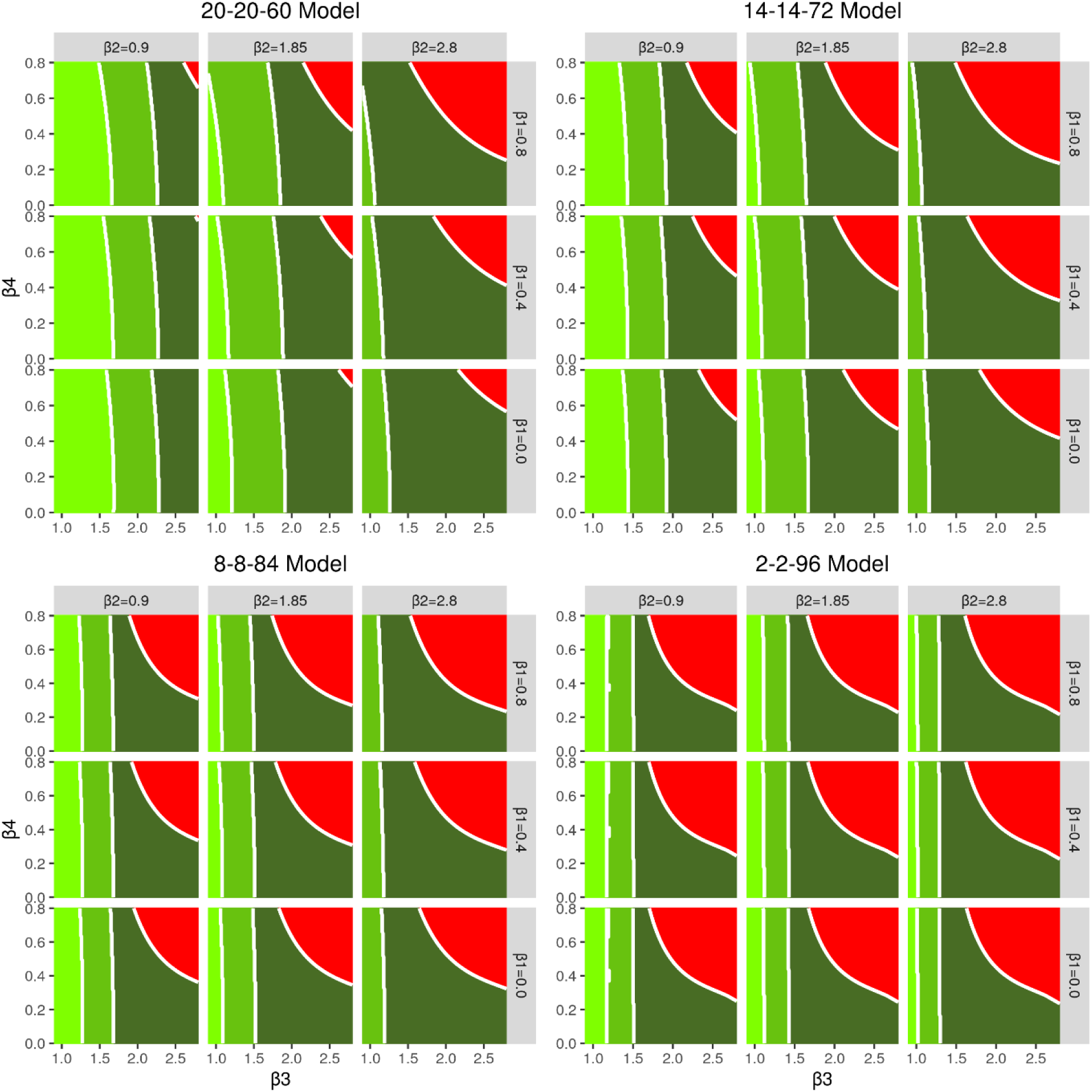
Heat maps showing the trade-off between relaxation (left to right on horizontal axis) and increasing protection (top to bottom on vertical axis) for the different models considered. The green shading indicates which of the policy objectives is met: Dark green: second peak of I_v_ is lower than the first peak. Middle green: as dark green plus all 2^nd^ peaks (I_v_, I_s_, I_g_) lower than 1^st^ peaks. Light green: As middle green but dI/dt is negative or zero for at least one year after the start of lockdown for all I-compartments. Red: none of the policy objectives are met.

## Discussion

We note several caveats to our findings. We used relatively simple models to explore a wide range of scenarios. These scenarios are not predictions; in our view there are too many uncertainties about the epidemiology of COVID-19 to make robust predictions beyond short-term projections of epidemic data. There are three important sources of uncertainty that may influence our results:

i. The contact structure between and within segments is not well quantified. We carried out an extensive sensitivity analysis (Figure 4) to identify critical elements of the contact matrix.
ii. Relaxing restrictions and increasing protection both involve changes in behaviour. These are difficult to predict in advance though they can be monitored in close to real time (Jarvis et al., 2020).
iii. Further, the relationships between behavioural changes and transmission rates are also difficult to predict so close monitoring of the epidemic remains essential.

Given these limitations, we simulated a range of plausible scenarios, consistent with available data. We find that a combination of increased protection of the vulnerable population and relaxation of restrictions (lockdown) on the non-vulnerable population can prevent an overwhelming second wave of the COVID-19 epidemic in the UK.

This result is driven by the build-up of population immunity during the first wave, particularly in the non-vulnerable population (Figure S1). The extent of population immunity for COVID-19 is uncertain (Kellam & Barclay, 2020). However, our analysis suggests that even short-lived population immunity will have a significant effect. It has been argued that short-lived immunity (average duration c. 1 year) will allow multiple waves of infection over many years (Kissler et al., 2020). In the absence of any acquired immunity to COVID-19 the epidemic becomes significantly more difficult to control (Figure S8).

Other key drivers are the size of the vulnerable population and their relative risk of severe infections. A smaller vulnerable population may be logistically easier to protect, and perhaps more likely to comply, but is likely to incur a smaller proportion of the severe disease burden. At the same time, a consequence of protecting a smaller proportion of the population and relaxing restrictions for a larger proportion is that overall transmission rates are higher. The implication is that S&S will be much more difficult to implement successfully if the proportion of the population designated vulnerable is too small. That said, as risk factors for severe COVID-19 infections become better understood it should be possible to define the vulnerable population more precisely.

Sensitivity analyses suggest that the most influential transmission rates are those between the vulnerable and general population segments (Figure 4). This is important because these rates can be reduced by physical distancing, which is considerably more difficult to do for the shielders. However, the same analysis also underlines the importance of transmission within the general population, which is the main reservoir of infection. It is therefore vital that transmission rates are kept as low as possible, even if this population is allowed to exit lockdown. Measures including self-isolation of cases, quarantining of affected households, contact tracing and voluntary physical distancing will be necessary to achieve this.

In all our scenarios the vulnerable segment is subject to increased protection indefinitely. S&S is also more likely to succeed if there is less or no relaxation of restrictions on shielders. These two observations underline the importance of both identifying the vulnerable and shielder populations as precisely as possible and of developing strategies for protection/shielding that minimise the disruption to normal activities, not least to ensure high levels of adherence.

Policy objectives also impact on the range of S&S strategies that could be used. The most restrictive policy objective we considered – not allowing any increase in the number of cases – cannot currently be achieved without physical distancing measures. This leaves very little room for relaxing lockdown measures even with greatly enhanced protection for the vulnerable.

A key component of S&S is behavioural modification, not only for the vulnerable and shielder segments but also for the general population. We note that appropriate advice could be issued quickly and cheaply, making this suitable for any country affected by COVID-19.

In addition, S&S could be greatly strengthened by infrastructure and technological support for effective biosecurity, both at institutional (e.g. care homes, hospitals) and household levels in order to keep transmission rates low between and within shielders and vulnerable populations. For maximum effectiveness biosecurity requires training, high standards of hygiene, effective personal protective equipment and screening of everyone in contact with the vulnerable population.

Intensive screening would, ideally, include daily checks for symptoms, daily tests for virus presence (preferably with results available the same day to prevent pre-symptomatic transmission), regular serological testing and monitoring of frequent contacts (e.g. household members) of shielders. If too large fraction of the population were to be classified as ‘shielders’ this would quickly overwhelm current testing capacity in the UK. Nonetheless, routine rapid testing of shielders could have a significant impact and further increase the scope for relaxing restrictions on the entire population (Figure S9).

Finally, we note that S&S would not be implemented in isolation. Measures such as contacting tracing (both traditional and app-based) could also facilitate exit from lockdown (Kucharski et al., 2020). In the long term effective therapeutics and vaccines may alleviate the need for restrictive physical distancing measures. Even then, however, we anticipate that COVID-19 biosecurity will need to be built into the daily routines and working practices of all hospitals, care homes, other vulnerable institutions and some households, affecting everyone who resides in, works in, or visits those locations.

## Data Availability

The data that support the findings of this study are openly available in the github repository at http://www.github.com/bvbunnik/COVID-19-enhanced-shielding.

## Acknowledgements

We are grateful to Wendy Barclay for helpful discussions and to Katie Atkins for a careful critique of a draft. We are grateful to the Royal Society’s Rapid Assistance in Modelling the Pandemic (RAMP) initiative for two anonymous reviews of the manuscript. All errors and omissions remain the responsibility of the authors. The views expressed in this paper are entirely the personal views of the authors.

